# Prediction and control of COVID-19 infection based on a hybrid intelligent model

**DOI:** 10.1101/2020.10.22.20218032

**Authors:** Gengpei Zhang, Xiongding Liu

## Abstract

The coronavirus (COVID-19) is a highly infectious disease that emerged in the late December 2019 in Wuhan, China, and it has caused a worldwide outbreak, which represents a major threat to global health. It is important to design prediction research and control strategies to crush its exploding. In this study, a hybrid intelligent model is proposed to simulate the spreading dynamics of COVID-19. First, considering the control measures, such as government investment, media publicity, medical treatment and law enforcement. The infection rates are optimized by genetic algorithm (GA), then a modified susceptible-infected-quarantined-recovered (SIQR) model is proposed, then the long short-term memory (LSTM) is imbedded into the SIQR model to design the hybrid intelligent model to further optimize other parameters of the system model to obtain the optimal predictive model and control measures. This study provide a reliable model to predict cases of infection and death, and reasonable suggestion to control COVID-19.

## Introduction

In the past six months, Chinese people have been on a high level of containment due to the outbreak of the coronavirus 2019 throughout China [1]. However, due to the convenient of global transportation, the virus has rapidly spread to all corners of the world. The coronavirus was first discovered in Wuhan, at the early stage, due to people lack of knowledge about the virus and the scarcity of medical resources, people were not aware of the virus. Relatively little knows about the disease, people neglect to control it in its early stages, until the earliest official announcement that the disease can be spread from person to person on January 20, 2020 [2]. Then, the government took a series of measures to prevent the spread of the disease on January 21, like Wuhan lockdown. Subsequently, some cities closured, with cities across the country sealing off. Governments took many measures by closing down public places, broadcasting propaganda, isolating people in their own homes, etc., which leads to the increased awareness of self-protection for people. The epidemic has been successful control in China, but now the world is not optimistic, with over 40 million people now cumulatively diagnosed worldwide. The cumulative number of deaths is over 1.2 million, especially in countries such as the United States, Brazil, Russia, and India, the pandemic is still quite severe today [3,4]. Therefore, each countries and states should adopt prevention and control strategies. Currently, it is very important to establish and analyze disease-spreading model to predict disease development trends in order to control and prevent the spread of the COVID-19.

Due to the outbreak of disease, vaccines and medicine treatments for the disease are still being researched, non-drug treatment becomes the main strategy to slow down the spread of the disease. The purpose of most of these prediction and control measures is to reduce the probability of infection spreading during direct or indirect contact [5]. For example, paying attention to personal hygiene and wearing a mask, keeping social distancing and closing some public space such as schools and workplaces, in order to decrease the opportunities of propagation from person to person. Other measures like cutting off the way of transmission of diseases, such as disinfection in public places, improving the level of sanitation in public places and so on. All of these measures can alleviate the propagation of the epidemic [6-9]. How to quantify the impact of various prevention and control strategies on disease spreading is of great significance to guide disease control.

A common method utilizes mathematical modelling to describe the spreading dynamics of infectious diseases, like Ebola, SARS. This can accurately describe the spread of disease among individuals in theoretical framework to guide the development of the prevention and control measures [10-13]. There are many researchers studied epidemic spreading have proposed some epidemic model and obtained some meaningful results. The classical epidemic models like susceptible-infected (SI), susceptible-infected-recovered (SIR), susceptible-exposed-infected-recovered (SEIR), susceptible-infected-quarantined-recovered-susceptible (SIQRS) [14-19]. Most of these models are applicable to describe disease spreading with a long incubation period, such as COVID-19. Based on data-driven, a modified SEIR model was proposed to analysis and prediction COVID-19 spread [20]. Considering the effect of quarantine, B. K. Mishra et.al proposed three quarantine models to analyze COVID-19 spreading [21]. Other recent studies, some researchers have considered the effects of the basic reproductive number, international conveyance, and some stochastic based regression models to prediction and control the COVID-19 disease [22,23]. However, traditional epidemic spreading models consider that all infected individual have the same infection rate, and the prediction of disease development trend has certain limitations. Although data-driven disease spreading models can accurately describe infection rates, the impact of government prevention and control measures on infection rates has not been quantitatively in detail. These measures, such as the laws, medical supplies, media coverage and investment, can reduce the spread of the disease. It is necessary to rationally arrange the optimal prevention and control strategies with limited resources to minimize the death rate.

To solve this problem, the GA and ANN hybrid method is proposed to optimize epidemic dynamics model and predict the COVID-19 spreading [24-27]. Genetic algorithm is an adaptive global optimization search algorithm formed by simulating the genetic and evolutionary process of biological species in the natural environment [28]. It uses the viewpoint of biogenetics and realizes the improvement of individual adaptability through the mechanism of natural selection, heredity and variation. Artificial intelligence (AI) is considered one of the most successful achievements of computer science, simulating the behavior of the human brain in data analysis. One of the AI branches is the artificial neural network (ANN). This information processing system, by a simulating strategy like communication between brain neurons, has become a tool for analyzing complex and real systems [29]. In recent years, ANN models have been developed to overcome the difficulties presented by health issues.

This article focuses on how to quantify the impact of the government’s prevention and control measures on the infection rate, then to obtain the optimal model of disease spreading and the most effective prevention and control strategy. Based on the proposed SIQR model, the hybrid artificial neural network (ANN) model embedded the genetic algorithm for predicting the COVID-19 in this article, and it introduces the important prediction and control strategies led by the government as well as the massive support participation from the public into the prediction calculation process. Furthermore, this article simulates the development of the epidemic based on the proposed hybrid prediction model and predicts the trend of the epidemic. Due to the fluctuation of virus detection capability, the epidemic data of China shows a sudden change on February 12 and 13, the data consistency is reduced. As a developing country, Brazil shows the epidemic data with periodicity and consistency, and then this article takes the Brazil data as analytic target. The simulation results based on the epidemic data of Brazil show that the proposed hybrid model could provide a basis for estimating the law of virus spread, and achieve accurate and robust performance. Moreover, the prediction results of our hybrid ANN-GN model is agree with the actual epidemic development trend, which demonstrates that the openness, transparency, and efficiency of data releasing are very important for establishing a modern epidemic prevention system.

The remainder of this article is organized as follows. Section “A hybrid model of COVID-19 spreading dynamics” introduces the framework of the proposed hybrid epidemic spreading model. Section “Methods” explains the method of GA and ANN to predict epidemic spreading. Section “Simulation results and discussion” provides the simulation results based on the epidemic data of Brazil and gives some discussion. The conclusions are provided at last.

### A hybrid model of COVID-19 spreading dynamics

In this section, we establish a mathematical model on COVID-19 based on some assumptions. There are some literatures proposed models mainly based on real clinical data, predict and control measures. In this paper, we modified the previous model proposed in [18], and then extend the model structure by designing different infection rate. The model network diagram and the interaction individual components demonstrated in Fig 1. Every individual in network can only exist one of four independent states, namely, susceptible, infected, quarantined and recovered. For simply, it can be denoted by S, I, Q, and R, respectively. Each link represents the transformation relationship between nodes. Here, infected individuals include symptomatic infected individuals and asymptomatic infected individuals. Susceptible individual is infected with probability m (M1, M2, M3 and M4) if it is connected to an infected individual. Infective individuals are quarantined with probability α. In the process of isolation, the asymptomatic infected individuals turn into symptomatic infected individuals with the probability ω. Quarantined individuals are treated with drugs that move into the recovered individuals with probability β. Some recovered individuals will relapse into infection due to their weakened immunity with probability γ. The probability of death during isolation is λ. Here, set a switch of city lockdown by the death rate, infection probability m is different based on the city situation (lockdown or not). The lockdown infection probability m is much lower for the strict government regulation.

**Fig 1.**
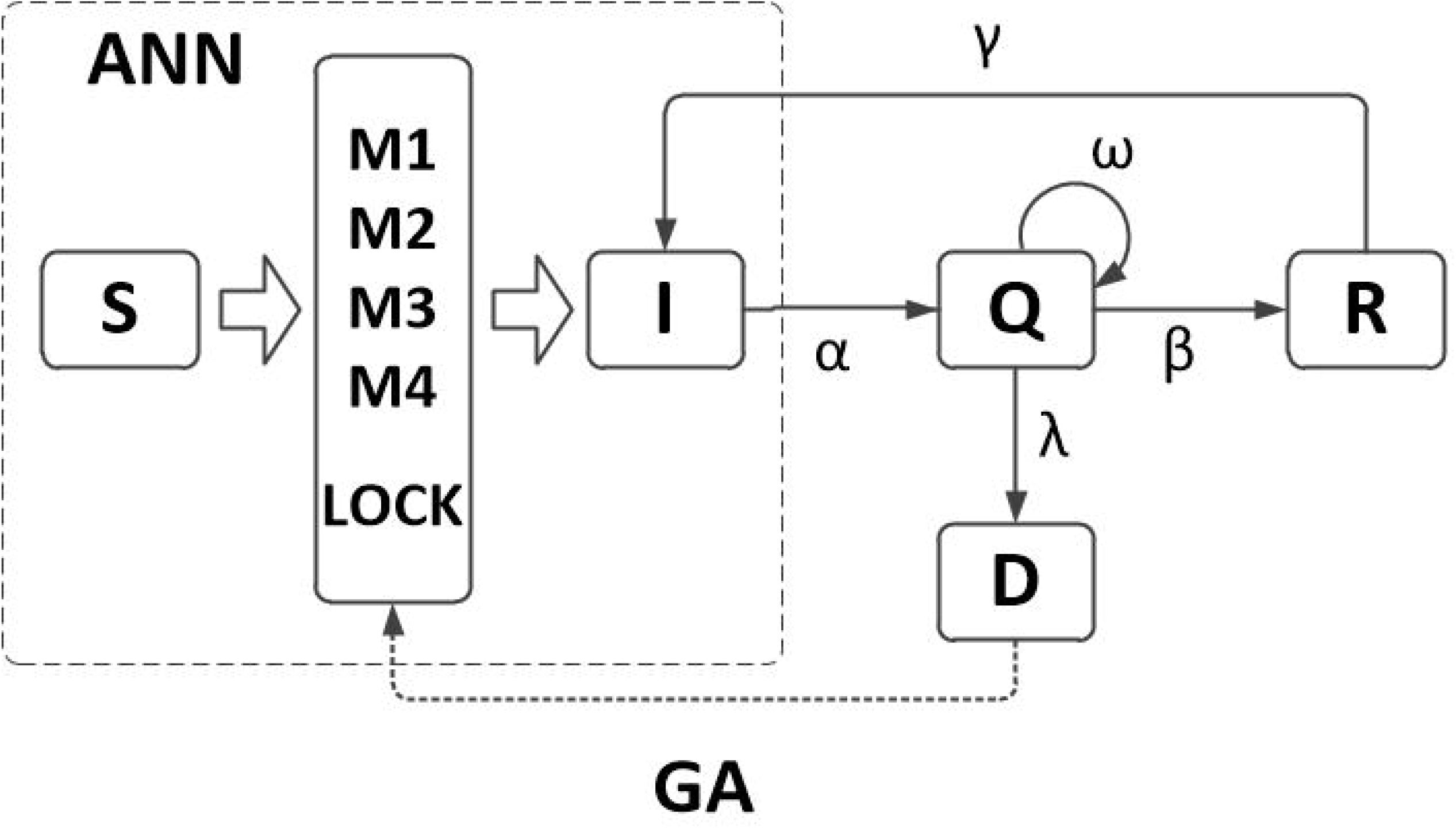
The flow diagram of the SIQR model.

According to the epidemic spreading, the dynamic equations can be written as follows:

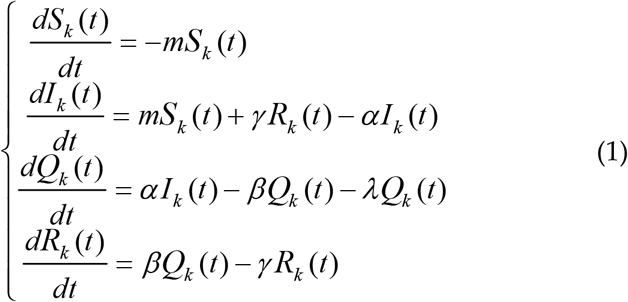

Using the normalization condition, the probability of death can be obtained:

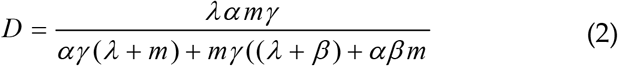

In this paper, the proportion of death is a very important parameter. It not only depicts the fatality rate of the disease in the process of transmission, but also reflects the effectiveness of prevention and control measures. For example, the money invested by the government can improve the medical level and effectively cut off the transmission route of the virus. Increasing publicity and awareness of prevention and control will also reduce the risk of infection. Law enforcement can also affect the spread of diseases, such as city closures and home quarantine. This paper argues that due to the strict control and isolation measures taken by the government during the epidemic, the infected cases cannot infect susceptible people after quarantine. However, asymptomatic infected persons also have some ability to transmit, and it is difficult to detect. Therefore, there is a certain relationship between the number of newly infected cases on Day t and the number of infected cases in the past k days. Moreover, the infection rate of patients is closely related to the time of infection. Since government measures can inhibit the spread of the disease, the infection rate of newly infected cases may vary from time to time on t day over the past k days. Further analyzing this difference and assigning different weights to different measures, we quantified the contribution of different measures to the infection rate at time t in newly infected cases. Then, the weighted accumulation was used to estimate the infection rate so as to establish the relevant epidemic air defense modeling.

In addition, in order to study the relationship between the prediction and control measures with infection rate of the SIQR epidemic spreading model, the method of GA and LSTM are used to optimize the spreading model. This paper considers the relationship between the rate of disease transmission and the measures taken by the government against the disease. The main factors include government investment, medical level, media publicity and law enforcement. Firstly, genetic algorithm is used to estimate the infection rate of the model, taking the acceptable mortality rate as fitness function and taking it as a basis for city closure parameters. At the same time, GA is further used to obtain the optimal means of government control by taking the minimum mortality rate as fitness function. The mutation law of GA is based on the interaction of four government measures. Furthermore, the infection rate bias and mortality bias were estimated through the LSTM network, the number of infected people was estimated by combining with the SIQR model, and the relevant parameters of the model were modified to obtain the best transmission model and predict the spread of the disease. By combining these two approaches, the optimal model of disease spreading and the optimal prevention and control strategies can be obtained, which can predict the number of infected and death cases based on the spreading model and development trend. The proposed framework is shown in Fig 2.

**Fig 2.**
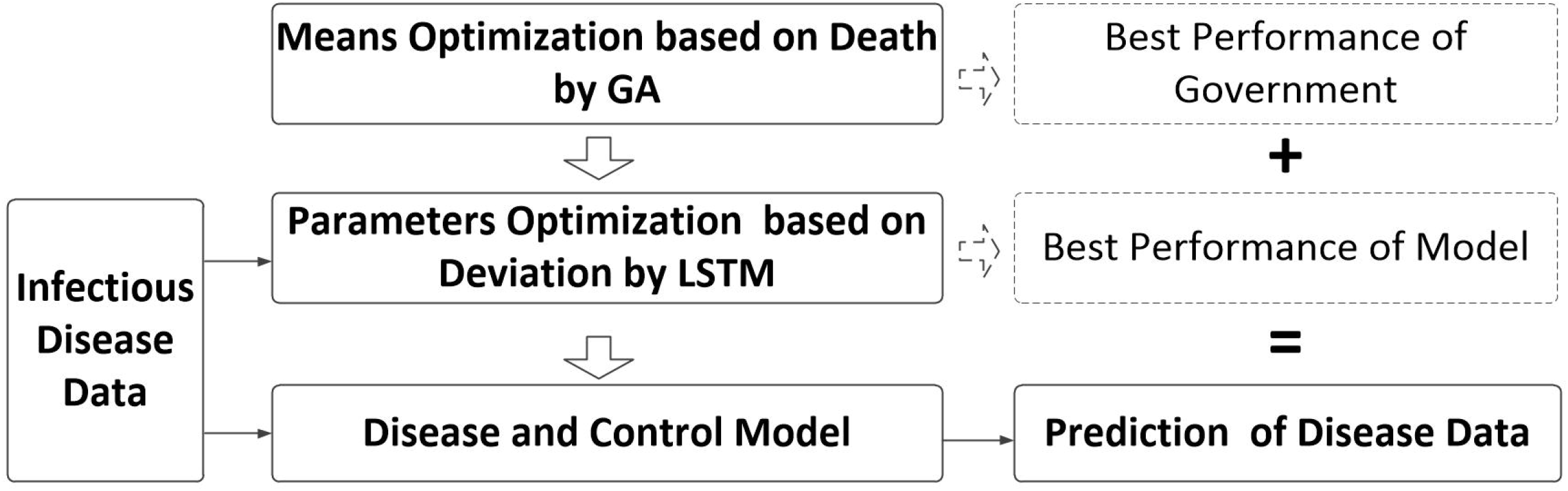
The hybrid model for COVID-19 prediction.

## Methods

Epidemiological investigation and modeling are effective tools to study the spread of epidemic diseases and have played their due roles in the prevention and control of public health events with global impact, such as SARS, MERS and H1N1 influenza. Many researchers have also achieved some results in novel coronavirus research using epidemiology and modeling analysis. The study of J. Chan et al. [30] found the first evidence of human-machine transmission. Then, some researchers combined with the analysis of some early cases and gave the mean incubation period and mean infection cycle of novel coronavirus. Traditional models of disease transmission believe that the number of new infections is related to the number of infected and vulnerable people, but these models lack in-depth analysis of parameters in the process of disease transmission. In the process of disease transmission, the implementation of different control measures has a great impact on the prevention and control of disease and the suppression of disease transmission. For example, the government’s publicity of disease prevention and control, the formulation of relevant laws and regulations, financial investment and other measures can affect the spread of disease. The study found significant differences in the rates of infection among people of different ages. The main purpose of this paper is to study the impact of government measures on the spread of disease and to minimize the mortality rate, and to consider the impact of different age groups on the spread of disease, finally, we can obtain the optimized measures and the best performance model to prediction, as showing in Fig 2.

Unlike the traditional SIQR epidemic spreading model, which consists of a single infected rate to describe the probability of infection. In this paper, we proposed a dynamic strategy to better represent the real world and the new COVID-19 disease. It mainly considers the impact of various measures taken by the government on the control of disease spreading. This paper considers the impact of government investment, media publicity, medical treatment and law enforcement on the rate of disease spreading, and its parameters are set as mi, i=1, 2, 3, 4, respectively. In addition, k_i_ is the weight of each measure. Thus, the rate of disease spreading can be written as:

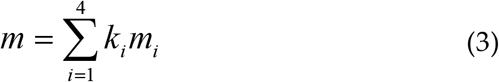

The GA is mainly used to optimize parameters of the model, it can be divided as two parts, and the first part describes that using the GA to optimize the conditions of city closure. First initialization model, set the initial value, with acceptable mortality as a condition of judgment. According to data analysis and relevant government regulations, a three-day mortality rate greater than 0.045 is defined here as an unacceptable mortality rate. When this unacceptable mortality rate is reached, will update and save the neural network parameters, as a basis for the sealing city.

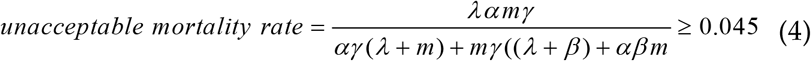

At the same time, further to run the model, with the minimum mortality as fitness function, further update neural network parameters by genetic algorithm.

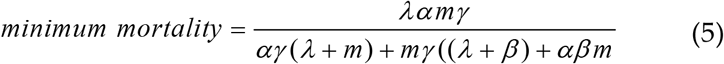

Using twice the genetic algorithm, the approximate optimized neural network parameters, namely the control measures taken by the government can be obtained. Further, the LSTM algorithm is used to modify other parameters of the model, and the historical data is compared with the data calculated by the model, such as the number of infections and deaths, to determine whether the system has reached the minimum error. The number of infections and deaths are further predicted. The flow chart of COVID-19 prediction algorithm is shown in Fig 3.

**Fig 3.**
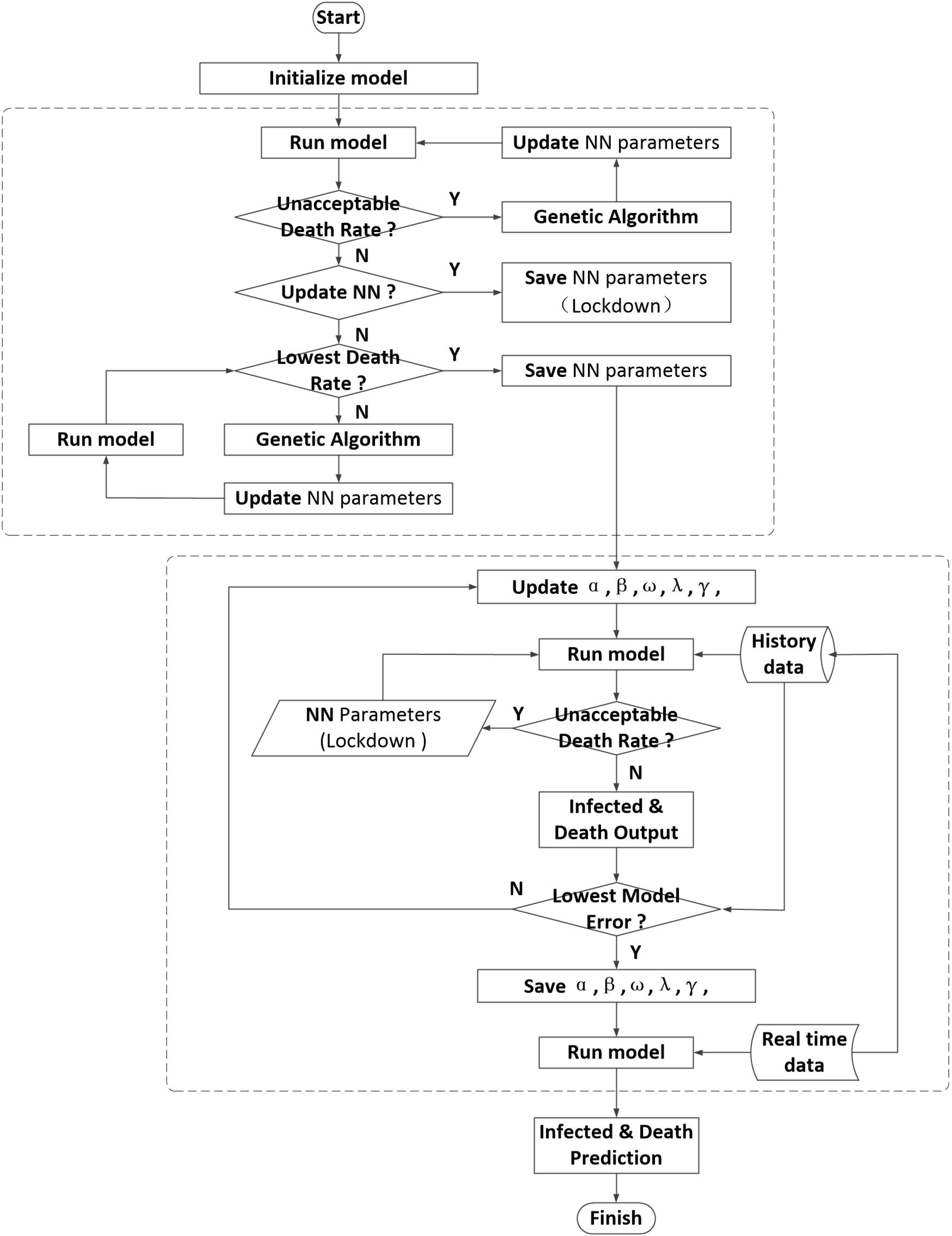
The flow chart of COVID-19 prediction algorithm.

New data on the daily increase in the number of infected cases can be obtained. Using the same method, it is also possible to process the number of new deaths per day.

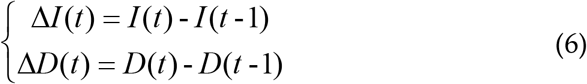

Here, *I(t)* is the cumulative number of infected cases in the previous t-day, *I(t-1)* is the cumulative number of infected cases in the previous t-day, and *ΔI* is the new number of infected cases in the first t-day. *D(t)* is the cumulative number of deaths in the first t-day, *D(t-1)* is the cumulative number of deaths in the first t-day, and *ΔD* is the additional number of deaths on the first t-day.

In order to get a more accurate model, the model parameters need to be further processed. The GA has optimized the weight parameters in the neural network, and other model parameters are optimized through the optimized neural network model. Here, LSTM neural network is used to optimize the model parameters. We collected historical data from February 26 to October 13 in Brazil, to train neural network. Here we discussed the model parameters through two judgments. First, when the mortality rate of the model reaches an unacceptable mortality rate, the neural network parameters of the closed city are used to process the system model parameters, and then the mortality rate and infection rate are further compared with the actual data. By obtaining the minimum model error, the optimized model parameters are finally obtained. Here, let the actual infected rate be ΔI and the infected rate under the regression exponential function be Δ I’, and use the neural network to predict the deviation between the actual mortality and the regressive infection rate. Similarly, the same approach can be applied to mortality and obtained ΔD and ΔD’. Assuming that B=ΔI’+ΔD’ is the deviation characteristic of the prediction, the LSTM method can be used to predict the model. The flow diagram of the prediction is shown in Fig 4.

**Fig 4.**
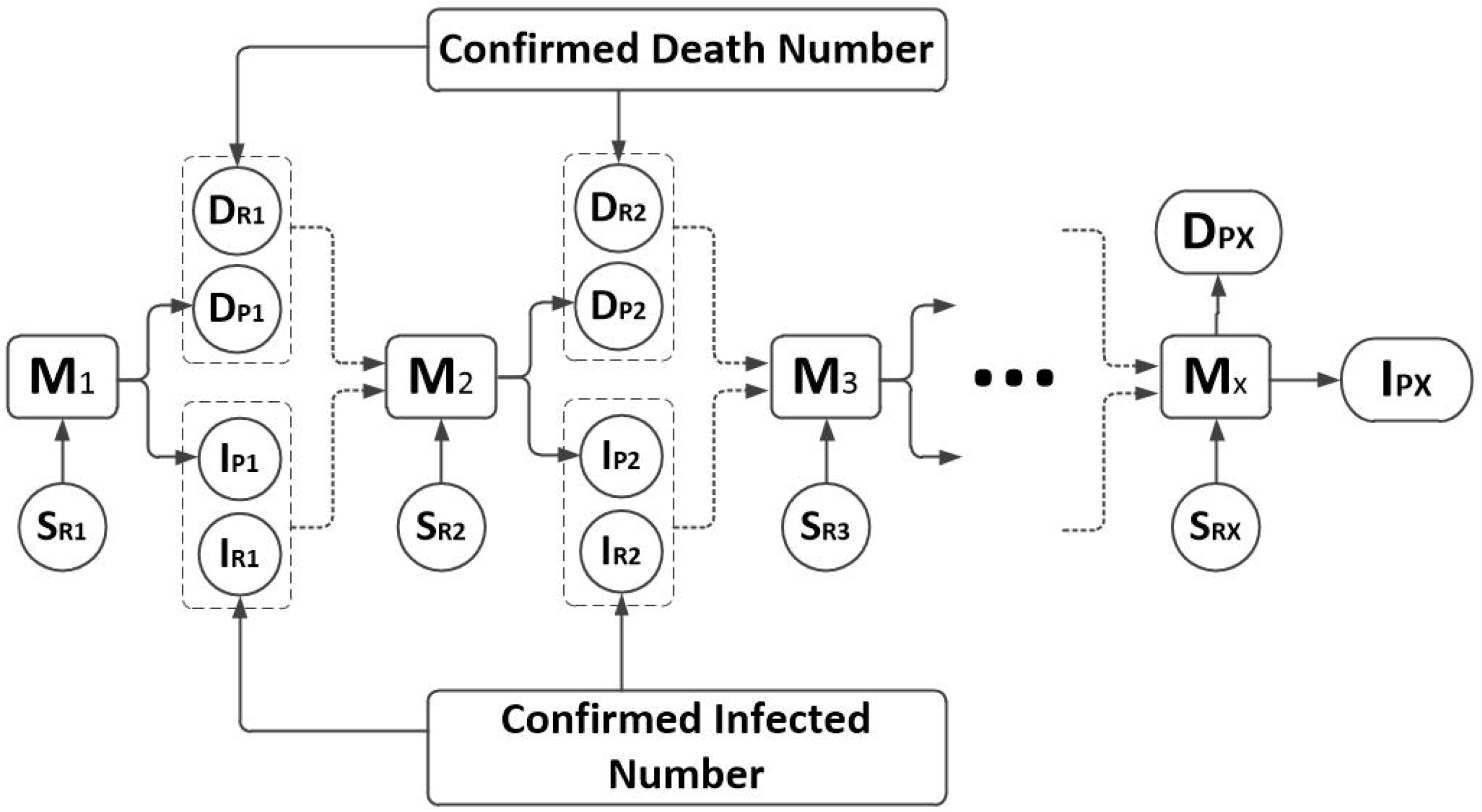
The LSTM method.

S_R1_ is the number of vulnerable infected persons on the first day, I_R1_ is the number of real-time infected cases on the first day, I_P1_ is the number of infected cases predicted on the first day, M_1_ is the initial model. Where S_RX_ is the number of vulnerable infected persons on day X, I_RX_ is the number of real-time infected cases on day X, I_PX_ is the predicted number of infected cases on day X, and M_X_ is the model on Day X.

## Simulation results and discussion

Based on GA in Fig 1, we computed the vulnerability coefficient k_i_ of four government measures. Without city lockdown, the infection probability m of government investment, media publicity, medical treatment and law enforcement are 0.174, 0.717, 0.021 and 0.853. With city lockdown, the infection probability m of government investment, media publicity, medical treatment and law enforcement are 0.085, 0.219, 0.107 and 0.349. There are two significant government measures media publicity and law enforcement, and the law enforcement is more important. Make a globally observation, each country invested huge amounts of money for COVID-19, and most medical workers tried their best to save lives. The difference of epidemic prevention results comes from the media publicity and law enforcement. Enhancing the influence of media can build a strong basis of epidemic prevention by offering the meaning of prevention measures. Considering the exception of inconsiderate and non-media audiences, the risk of disease spreading still exist. The efficiency of law enforcement is the crucial insurance to save the effort of medical workers.

We simulated the disease development and analyze the output of the model in 231 days with real epidemic data available from Brazil (February 26 to October 13). Fig 5a represents the number of predicted and confirmed per days, the model predicts infected per days of the last 17 days before October 13. Considering the lockdown from March 21 to July 31, the government realized the necessity of quarantine. From the data trend of infection confirmed per days over this period, the law enforcement shows insufficient efficiency. Supposing the government gave the best performance to control the disease, we performed the simulation about the development of infection and death per days. Fig 5b is the infected prediction with best performance of government; the possible infected number would much lower with effective government effort. With the comparison between Fig 5a and Fig 5b, it is obvious that there is a great impact on the prediction in the middle and later period due to the large changes in the data after government make best moves. Fig 5c is the daily-infected prediction error of model for last 17 days. Considering the enormousness of infection confirmed number, Fig 5c shows the accuracy of model prediction without best performance of government.

**Fig 5.**
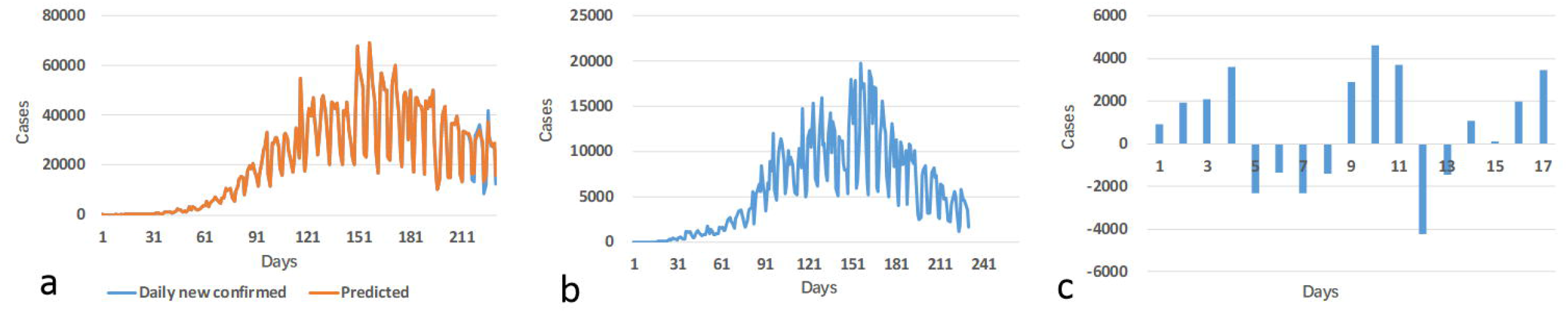
Brazil infected cases. a) Infection of predicted and confirmed per days, b) Infection prediction with best government performance, c) Infection prediction error.

To better understand the model performance, we simulated deaths in Brazil. Fig 6a represents the number of new death cases per day, this indicates that at the beginning of the epidemic, due to limited understanding of disease transmission and limited detection efforts, there will be omissions in disease statistics, leading to a similar increase trend with the infected cases. Meanwhile, Fig 6a represents the new predicted death cases per days of the last 17 days before October 13, which shows the difference with daily new death cases. Fig 6b is the daily death prediction with best performance of government. While the continuous improvement of detection methods and the continuous promotion of detection scope will be carried out, the government could strengthen the control of the disease, so the daily death cases would gradually decrease. Fig 6c is the daily death prediction error of model for last 17 days. Combine Fig 6a, we can see that the LSTM method have a good performance to predict COVID-19 disease. The method can predict the trend of the disease over a longer period with the performance of government.

**Fig 6.**
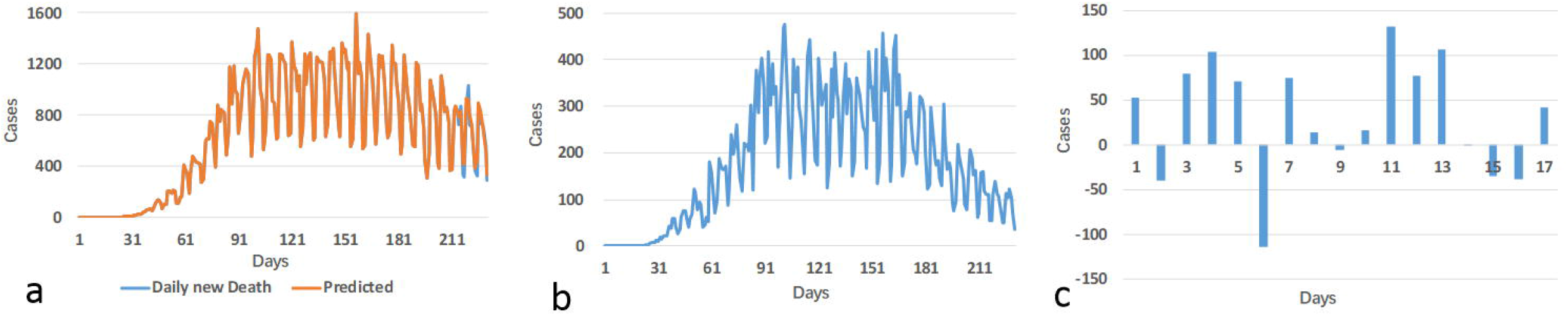
Brazil death cases. a) Death of prediction and confirmed per days, b) Death prediction with best government performance, c) Death prediction error.

## Conclusions

Based on the SIQR disease-spreading model, this paper seeks the best government performance from the four aspects by GA; and then proposed a hybrid prediction model with LSTM network. By analyzing Brazil data from February 26 to October 13, we analyzed new infected cases, new death cases per day. It is found that media publicity and law enforcement have more contribution to reduce transmission rate. With best government performance, the tread of COVID-19 in Brazil could make a difference. The prediction results of this model are highly consistent with the actual epidemic situation, which proves that the hybrid model proposed in this paper can efficiently analyze the transmission law and development trend of the virus.

## Data Availability

All data, models, and code generated or used during the study appear in the submitted article.

## References

1. Huang C, Wang Y, Li X, Ren L, Zhao J, et al. (2020) Clinical features of patients infected with 2019 novel coronavirus in Wuhan, China. Lancet 395: 497–506. https://doi.org/10.1016/S0140-6736(20)30183-5

2. Ye F, Xu S, Rong Z, Xu R, Liu X, et al. (2020) Delivery of infection from asymptomatic carriers of COVID-19 in a familial cluster. Int J Infect Dis 94: 133–138. https://doi.org/10.1016/j.ijid.2020.03.042

3. Crokidakis N (2020) Modeling the early evolution of the COVID-19 in Brazil: Results from a Susceptible–Infectious–Quarantined–Recovered (SIQR) model. Int J Mod Phys C: 2050135-2050144. https://doi.org/10.1142/s0129183120501351

4. Bhatraju PK GB, Nichols M, Kim R, Jerome KR, Nalla AK, Greninger AL, Pipavath S, Wurfel MM, Evans L, Kritek PA, West TE, Luks A, Gerbino A, Dale CR, Goldman JD, O’Mahony S, Mikacenic C. (2020) Covid-19 in Critically Ill Patients in the Seattle Region - Case Series. New Engl J Med 382: 2012–2022. https://doi.org/10.1056/NEJMoa2004500

5. Bai Y, Yao L, Wei T, Tian F, Jin D-Y, et al. (2020) Presumed Asymptomatic Carrier Transmission of COVID-19. JAMA 323: 1406–1407. https://doi.org/10.1001/jama.2020.2565

6. Lai C-C, Liu YH, Wang C-Y, Wang Y-H, Hsueh S-C, et al. (2020) Asymptomatic carrier state, acute respiratory disease, and pneumonia due to severe acute respiratory syndrome coronavirus 2 (SARS-CoV-2): Facts and myths. J Microbiol Immunol 53: 404–412. https://doi.org/10.1016/j.jmii.2020.02.012

7. Lan L XD, Ye G, Xia C, Wang S, Li Y, Xu H. (2020) Positive RT-PCR Test Results in Patients Recovered From COVID-19. JAMA 323: 1502–1503. https://doi.org/10.1001/jama.2020.2783

8. Lauer SA, Grantz KH, Bi Q, Jones FK, Zheng Q, et al. (2020) The Incubation Period of Coronavirus Disease 2019 (COVID-19) From Publicly Reported Confirmed Cases: Estimation and Application. Ann Intern Med 172: 577–582. https://doi.org/10.7326/M20-0504

9. Linton N, Kobayashi T, Yang Y, Hayashi K, Akhmetzhanov A, et al. (2020) Incubation Period and Other Epidemiological Characteristics of 2019 Novel Coronavirus Infections with Right Truncation: A Statistical Analysis of Publicly Available Case Data. J Clin Med 9: 538. https://doi.org/10.1101/2020.01.26.20018754

10. Zhou Y, Ma Z, Brauer F (2004) A discrete epidemic model for SARS transmission and control in China. Math Comp Model 40: 1491–1506. https://doi.org/10.1016/j.mcm.2005.01.007

11. Poletto C, Pelat C, Levy-Bruhl D, Yazdanpanah Y, Boelle PY, et al. (2014) Assessment of the Middle East respiratory syndrome coronavirus (MERS-CoV) epidemic in the Middle East and risk of international spread using a novel maximum likelihood analysis approach. Euro SurvII 19: 20824. https://doi.org/10.2807/1560-7917.es2014.19.23.20824

12. Zhang Q, Sun K, Chinazzi M, Pastore Y Piontti A, Dean NE, et al. (2017) Spread of Zika virus in the Americas. Proc Natl Acad Sci 114: 4334–4343. https://doi.org/10.1073/pnas.1620161114

13. Nishiura H (2011) Real-time forecasting of an epidemic using a discrete time stochastic model: a case study of pandemic influenza (H1N1-2009). Bio Med Eng OnLine 10: 15. https://doi.org/10.1186/1475-925x-10-15

14. Yang M, Chen G, Fu X (2011) A modified SIS model with an infective medium on complex networks and its global stability. Physica A 390: 2408–2413. https://doi.org/10.1016/j.physa.2011.02.007

15. Li T, Liu X, Wu J, Wan C, Guan Z-H, et al. (2016) An epidemic spreading model on adaptive scale-free networks with feedback mechanism. Physica A 450: 649–656. https://doi.org/10.1016/j.physa.2016.01.045

16. Daley DJ, Kendall DG (1964) EPIDEMICS AND RUMOURS. Nature 204: 1118. https://doi.org/10.1038/2041118a0

17. Zhu G, Fu X, Chen G (2012) Spreading dynamics and global stability of a generalized epidemic model on complex heterogeneous networks. Appl Math Model 36: 5808–5817. https://doi.org/10.1016/j.apm.2012.01.023

18. Li T, Wang Y, Guan Z-H (2014) Spreading dynamics of a SIQRS epidemic model on scale-free networks. Commun Nonlinear Sci Numer Simul 19: 686–692. https://doi.org/10.1016/j.cnsns.2013.07.010

19. Zhang J, Sun J (2014) Stability analysis of an SIS epidemic model with feedback mechanism on networks. Physica A 394: 24–32. https://doi.org/10.1016/j.physa.2013.09.058

20. Zhang Y, Yu X, Sun H, Tick GR, Wei W, et al. (2020) Applicability of time fractional derivative models for simulating the dynamics and mitigation scenarios of COVID-19. Chaos Soliton Fract 138: 109959. https://doi.org/10.1016/j.chaos.2020.109959

21. Mishra BK, Keshri AK, Rao YS, Mishra BK, Mahato B, et al. (2020) COVID-19 created chaos across the globe: Three novel quarantine epidemic models. Chaos Soliton Fract 138: 109928. https://doi.org/10.1016/j.chaos.2020.109928

22. Alkahtani BST, Alzaid SS (2020) A novel mathematics model of covid-19 with fractional derivative. Stability and numerical analysis. Chaos Soliton Fract 138: 110006. https://doi.org/10.1016/j.chaos.2020.110006

23. Mandal M, Jana S, Nandi SK, Khatua A, Adak S, et al. (2020) A model based study on the dynamics of COVID-19: Prediction and control. Chaos, Solitons & Fractals 136: 109889. https://doi.org/10.1016/j.chaos.2020.109889

24. Car Z, Baressi Šegota S, Anđelić N, Lorencin I, Mrzljak V (2020) Modeling the Spread of COVID-19 Infection Using a Multilayer Perceptron. Comput Math Method M 2020: 5714714. https://doi.org/10.1155/2020/5714714

25. Arora P, Kumar H, Panigrahi BK (2020) Prediction and analysis of COVID-19 positive cases using deep learning models: A descriptive case study of India. Chaos Soliton Fract 139: 110017. https://doi.org/10.1016/j.chaos.2020.110017

26. Haghshenas SS, Pirouz B, Piro P, Na KS, Cho SE, et al. (2020) Prioritizing and Analyzing the Role of Climate and Urban Parameters in the Confirmed Cases of COVID-19 Based on Artificial Intelligence Applications. Int J Environ Res Public Health 17: 3730. https://doi.org/10.3390/ijerph17103730

27. Wu X, Hui H, Niu M, Li L, Wang L, et al. (2020) Deep learning-based multi-view fusion model for screening 2019 novel coronavirus pneumonia: A multicentre study. Eur J Radiol 128: 109041. https://doi.org/10.1016/j.ejrad.2020.109041

28. Miralles-Pechuán L, Jiménez F, Ponce H, Martínez-Villaseñor L (2020) A Deep Q-learning/genetic Algorithms Based Novel Methodology For Optimizing Covid-19 Pandemic Government Actions. arXiv e-prints: 2005.07656.

29. Wong Z, Zhou J, Zhang Q (2018) Artificial Intelligence for infectious disease Big Data Analytics. Infect Dis Health 24: 44–48. https://doi.org/10.1016/j.idh.2018.10.002

30. Chan JF-W, Yuan S, Kok K-H, To KK-W, Chu H, et al. (2020) A familial cluster of pneumonia associated with the 2019 novel coronavirus indicating person-to-person transmission: a study of a family cluster. Lancet 395: 514–523. https://doi.org/10.1016/S0140-6736(20)30154-9

